# Characteristics of the sexual networks of gay, bisexual, and other men who have sex with men in Montréal, Toronto, and Vancouver: implications for the transmission and control of mpox in Canada

**DOI:** 10.1101/2023.08.31.23294912

**Authors:** Fanyu Xiu, Jorge Luis Flores Anato, Joseph Cox, Daniel Grace, Trevor A. Hart, Shayna Skakoon-Sparling, Milada Dvorakova, Jesse Knight, Linwei Wang, Oliver Gatalo, Evan Campbell, Terri Zhang, Hind Sbihi, Michael A. Irvine, Sharmistha Mishra, Mathieu Maheu-Giroux

## Abstract

**Background:** The 2022-2023 global mpox outbreak disproportionately affected gay, bisexual, and other men who have sex with men (GBM). In Canada, >70% of cases thus far have been among GBM in Montréal, Toronto, and Vancouver. We examined how the distributions of sexual partners 1) varied by city and over time related to the COVID-19 pandemic and 2) were associated with mpox transmission.

**Methods:** The *Engage Cohort Study* (2017-2023) recruited GBM via respondent-driven sampling in Montréal, Toronto, and Vancouver (n=2,449). We compared numbers of sexual partners in the past 6 months across cities and three time periods: pre-COVID-19 pandemic (2017-2019), pandemic (2020-2021), and post-restrictions (2021-2023). We modeled the distribution of sexual partner numbers using Bayesian negative binomial regressions and post-stratification, adjusting for sampling design and attrition. We estimated the basic reproduction number (*R_0_*), secondary attack rate (SAR), and cumulative incidence proportion of mpox using the fitted distributions and case timeseries.

**Results:** The pre-COVID-19 pandemic distribution of sexual partner numbers was similar across cities: participants’ mean number of partners was 10.3 (95%CrI: 9.3-11.3) in Montréal, 12.8 (11.1-14.7) in Toronto, and 10.6 (9.41-11.9) in Vancouver. Partner numbers decreased during the pandemic in all cities. Post-restrictions, sexual activity increased but remained well below pre-pandemic levels. Based on reported cases and post-restrictions distributions, the estimated *R_0_* (2.4-2.6) and cumulative incidences (0.6-0.9%) were similar across cities. The estimated average SAR across cities was 79%.

**Conclusion:** GBM sexual activity after restrictions were lifted remained below pre-pandemic levels. Comparable sexual partner distributions across cities may explain similarities in mpox *R_0_* and cumulative incidence across cities. Public health authorities should consider the risk of mpox resurgence for future vaccination and surveillance strategies as sexual activity is expected to recover.

## Background

A global outbreak of mpox (the disease caused by monkeypox virus) unfolded from May–October 2022, predominantly affecting gay, bisexual, and other men who have sex with men (GBM). The outbreak was unprecedented in its spread through sexual networks, number of cases generated, and geographical distribution, with most of the nearly 90,000 confirmed cases worldwide (May 2022–June 2023) occurring among GBM in regions with no previous history of reported transmission.^1–3^ These unusual transmission patterns of mpox virus were recognized by the *World Health Organization* (WHO) as a public health emergency of international concern (lasting from July 2022 to May 2023).^4^

In Canada, over 70% of reported cases were concentrated among GBM in the three largest cities: Montréal, Toronto, and Vancouver.^5–7^ Mpox cases were identified at the frontlines, in GBM sexual health clinics, with swift responses from community, clinical, and public health partners.^6,7^ Since December 2022, sporadic new cases have been reported to the *Public Health Agency of Canada*,^8^ and questions remain regarding the future risk of mpox resurgence.^9^

An important factor shaping transmission during the 2022–2023 mpox outbreak was the structure of GBM sexual networks.^10,11^ Studies from Europe and North America attributed local outbreaks to densely clustered sexual networks among GBM with a high number of sexual partners.^12–15^ Additionally, earlier case investigations revealed close linkages to international travel and sex-on-premises venues.^2,12,16^ The concept of “core group” in sexually transmitted infections posits that a small group of individuals with a high number of sexual partners disproportionally contribute to transmission.^17^ It recognizes that heterogeneity in sexual partners is crucial to transmission dynamics. In other words, the average number of sexual partners from a chosen member of the sexual network (i.e., degree) is not as informative as the distribution of sexual partner numbers (i.e., *degree distribution*) of that network. As such, mathematical modeling suggested that the basic reproduction number (*R_0_*) of mpox —the expected number of secondary cases arising from an initial infection in an entirely susceptible population— may be significantly greater than 1 as reported among GBM in the United Kingdom.^14^ Estimates of *R_0_*from other modeling studies based on European and Canadian populations ranged from 1.5 to 4.3.^18^

Although these findings have provided insights into the transmission dynamics of mpox, there remains uncertainty regarding how GBM sexual networks in major Canadian cities shaped transmission. The outbreak occurred in the aftermath of the COVID-19 pandemic, which may have affected usual sexual networks of GBM. For instance, the lifting of travel restrictions and other public health measures may have increased the number and types of sexual partnerships formed and facilitated international dissemination of the virus.^12,16,19,20^

Given these uncertainties, we leveraged data from the *Engage Cohort Study* and public mpox incidence data to improve our understanding of the relationship between sexual network density and mpox transmission during 2022–2023 in Montréal, Toronto, and Vancouver. Specifically, we estimated the distribution of sexual partner numbers among GBM in each city, and investigated how these distributions changed over time to assess the influence of COVID-19 pandemic on sexual behaviours. We also assessed the transmission potential of mpox in each city by estimating the *R_0_* and cumulative mpox incidence in each city, and the average secondary attack rate (SAR) across cities.

## Methods

### Study setting and population

The *Engage Cohort Study* (*Engage*; 2017–present) is a prospective, population-based cohort study of GBM in Montréal, Toronto, and Vancouver. Eligible participants were self-identified cis or trans men living in one of the three cities, aged ≥16 years, who reported sex with another man in the past 6 months (P6M), understood English or French, and provided written consent. From February 2017 to August 2019, participants were recruited using respondent-driven sampling (RDS), a method used to sample hard-to-reach populations to estimate more representative population characteristics.^21^ Initial participants were purposively selected to represent diverse characteristics of the GBM community, and all participants were invited to recruit up to six peers in their social networks. Details of the cohort, including use of the RDS method, have been detailed in previous research.^22,23^

To investigate changes in sexual behaviours related to the COVID-19 pandemic, we used data from baseline visits and two follow-up visits:

- The *pre-pandemic period* was defined as the participants’ baseline visit (February 2017– August 2019).
- The *pandemic period* was the earliest follow-up visit that occurred between June 2020– November 2021, covering successive waves of COVID-19 pandemic measures. The start of this period was chosen because *Engage* study visits only resumed three months after the COVID-19 pandemic was declared by the WHO (March 11th, 2020)^24^ . The end of the period was the start of the post-restrictions period.
- The *post-restrictions period* was defined as the latest follow-up visit that occurred between December 2021–February 2023. To ensure consistency of the time periods across cities, we defined the start of this period based on the easing of entry requirements for non-essential travel into Canada (September 3, 2021).^25^ The start of the time period was shifted forward by three months to account for the 6-month recall period used in the *Engage* questionnaire. Additionally, we used travel restrictions given the history of international travel reported from initial mpox case investigations. The end of this period was the latest available data cut from the *Engage Cohort Study* (February 2023).

### Variables

Our primary outcome was the self-reported number of sexual partners in the P6M, measured through the question “*During the PAST 6 MONTHS, with how many guys have you had any kind of sex (anal, oral, mutual masturbation, rimming, frontal/vaginal, etc.)?*”. Informed by epidemiological data on mpox cases,^2,12,26^ the following bio-behavioral variables were considered correlates of the number of sexual partners in the P6M in the analyses (details in ***Table S1***):

- Age (18-29, 30-39, 40-49, 50-59, ≥60 years);
- Relationship status and sexual arrangement (no relationship, exclusive relationship, open relationship, unclear);
- HIV status (determined using 4th generation testing with a confirmatory assay; or self-report if testing data unavailable [4% participants at baseline]);
- Visit to bathhouses and/or sex clubs at least once in the P6M (binary);
- Attendance of group sex events at least once in the P6M (binary);
- Use of dating apps to find partners at least once in the P6M (binary); and
- Participation in transactional sex at least once in the P6M (i.e., receive money and/or goods in exchange for sex; binary).

Missing values for the last four variables were handled using the missing indicator method for all analyses.^27^

### Distribution of sexual partner numbers, rationale for weighting, and computation of weights

To estimate the distribution of sexual partner numbers in each city, we modeled the observed distributions in a Bayesian framework. Briefly, we first fitted a negative binomial regression model to the number of sexual partners in P6M, using the correlates mentioned above as covariates in the model. We then incorporated sampling and attrition weights via post-stratification, by using the fitted posterior sample of the number of sexual partners in P6M for each participant. Lastly, we computed the fitted population distribution of sexual partner numbers based on the post-stratified samples.

We chose a regression approach over direct distribution fitting to examine individual correlates and to incorporate survey sampling and attrition weights. Regression models were fitted using Hamiltonian Monte Carlo in *Stan*, with 4,000 iterations (2 chains, 2,000 burn-in iterations, no thinning), using non-informative priors and assessing convergence via traceplots and *R̂*. We then used the posterior distribution of each participant’s outcome to estimate the population distribution of sexual partner numbers, incorporating RDS-II weights and inverse probability of censoring weights (IPCW) via post-stratification. For the pre-COVID-19 pandemic time period, only RDS-II weights were used. To adjust for attrition in the pandemic and post-restrictions time periods, we used the product of RDS-II weights and IPCW (henceforth RDS-IPC weights).

Given that *Engage* is an RDS sample, we used RDS-II weights to ensure the estimated distributions of sexual partner numbers were more representative of the target population (sexually active GBM in each city). In RDS samples, participants with larger social networks have a higher chance of being recruited; RDS-II weights adjust for this sampling design by assigning a weight that is inversely proportional to the self-reported network size.^21^ To adjust for attrition at follow-up visits, we used IPCWs to reduce potential biases stemming from correlation between the outcome (number of sexual partners in P6M) and being lost to follow-up.^28^

Finally, to compare the distribution of sexual partner numbers between a pair of cities or time periods, we computed the proportion of the iterations (posterior distribution samples) that had greater cumulative density for ≥25 and ≥100 partners. These thresholds were based on a previous modeling study from the Netherlands, which estimated that GBM with a mean of 25 partners in the P6M accounted for about 20% of mpox cases, and those with a mean of about 100 partners accounted for over 60% of cases.^29^

### Basic reproduction numbers, secondary attack rate, and cumulative incidence proportion

The basic reproduction number *R_0_* is a measure of an infectious agent’s transmission potential within a given network.^30,31^ We first estimated the *R_0_* of mpox in each city based on the fitted distribution of GBM’s sexual partner numbers in the P6M (post-restrictions period), by using the next-generation matrix (NGM) approach.^32^ Briefly, we partitioned the GBM population in each city into 100 sexual activity groups based on the number of sexual partners from the RDS-IPC-weighted posterior distribution. For each group, we computed the size (i.e., proportion of GBM in the group) and the average number of sexual partners. We constructed the transmission, transition and auxiliary matrices based on a Susceptible-Exposed-Infectious-Removed model, assuming no interventions or behaviour changes during the analysis period. The three matrices allowed for the construction of a NGM, and *R_0_* was computed as the largest non-zero eigenvalue of this matrix. We repeated the procedure for each city at the post-restrictions period under a range of SAR^33^, defined here as the proportion of sexual partnerships that could be expected to result in transmission from an infected individual.^34^

Given the uncertainty in the SAR of mpox, we also estimated *R_0_*from the reported mpox cases from Québec, Ontario, and British Columbia, where Montréal, Toronto, and Vancouver are located, respectively. Province-wide mpox incidence was assumed to reflect transmission in each city given that the majority of the cases were reported from these cities.^5–7^ Briefly, we used publicly available data on reported mpox cases^6^ to estimate this *R_0_* using the formula *R*_0_ =: 1 + *D*Λ, where the duration of infectiousness D is assumed to be 17.3 days,^35–38^ and the epidemic growth rate Λ was estimated from the slope of the log cumulative cases over time. We used the first 50 days after the first case in each province, which corresponded to the period of initial exponential growth.^33^ We then compared the estimates of *R_0_* calculated from the next-generation matrix to the case series to estimate the SAR. We estimated the SAR as the SAR value necessary for the two methods of calculating *R_0_* to be equal. Lastly, to quantify potential increases in mpox transmission if sexual activity recovers to pre-COVID-19 levels, we estimated the *R_0_* using the NGM method based on the pre-pandemic distribution of sexual partner numbers and the average SAR across cities.

Lastly, we estimated the cumulative incidence proportion of mpox cases among GBM during the 2022–2023 outbreak in each city using GBM population size estimates from previous studies. Briefly, we assumed that sexually active GBM (sex with a man in P6M) constituted 2.9% of the ≥15-year-old male population in each city.^39–41^ For each city’s population size, we used the 2021 census population counts of the corresponding census metropolitan area.^42^

### Sensitivity analyses

We performed three sensitivity analyses. First, to verify the robustness of results regarding the distribution of sexual partner numbers to our weighting approach, we repeated the analyses restricting the analytical sample to participants who had visits at all three timepoints. Second, as age is an important determinant of sexual activity and differences in the age distribution of the participants across the three cities have been reported,^43^ we performed regression-based standardization. Using all model covariates, we standardized the distributions to the covariate distribution observed in Vancouver. Lastly, given uncertainty regarding mpox transmission probabilities through different types of sex acts,^2,44^ we repeated the analyses focusing on the number of anal sex partners as the outcome.

All analyses were conducted using *R* (version 4.2.2) and *RStan* (version 2.21.7). Additional details on methods can be found in the *Supplementary Methods*. The code used for analyses is available from GitHub (https://github.com/pop-health-mod/mpox-engage-sex-networks).

### Ethics

Ethics approval was obtained from the Research Institute of the McGill University Health Centre and the Research Ethics Office of the Faculty of Medicine and Health Sciences, McGill University (A06-M32-23B), Toronto Metropolitan University (REB #2016-113), the University of Toronto (protocol #00033527), St. Michael’s Hospital (REB #17-043), the University of Windsor (REB #33443), the University of British Columbia (H16-01226), Providence Health Care (H16-01226), the University of Victoria (H16-01226), Simon Fraser University (H16-01226).

## Results

### Study population

There were 2,449 GBM recruited to *Engage*, with 1,179 participants in Montréal, 517 in Toronto, and 753 in Vancouver. Retention over the study period was slightly higher in Montréal, where 70% and 67% of participants had at least one visit during the pandemic and post-restrictions period, respectively, compared to 58% and 56% for Toronto, and 60% and 52% for Vancouver (***Table S2***).

Accounting for RDS-II, participants in Montréal were older on average than in Toronto and Vancouver—37% aged ≥40 years, versus 25% and 29%, respectively. In all three cities, approximately half of participants reported not being in a relationship at baseline. There were fewer participants living with HIV in Montréal (14%; 95% Confidence Interval [CI]: 12%–16%), compared to 22% (95%CI: 19%–26%) in Toronto and 20% (95%CI: 18%–23%) in Vancouver. More participants reported attending bathhouses and group sex in Toronto (39% [95%CI: 35%– 43%] and 23% [95%CI: 19%–26%], respectively) than in Montréal (31% [95%CI: 29%–34%] and 16% [95%CI: 14%–18%]) and Vancouver (29% [95%CI: 26%–32%] and 21% [95%CI: 18%–24%]). Lastly, the RDS-II-weighted mean number of sexual partners in the P6M was 8.7 (95%CI: 7.4–9.9) in Toronto, compared to 8.1 (95%CI: 6.9–9.4) in Montréal and 8.0 (95%CI: 6.8–9.2) in Vancouver (***Table 1***).

**Table 1.**
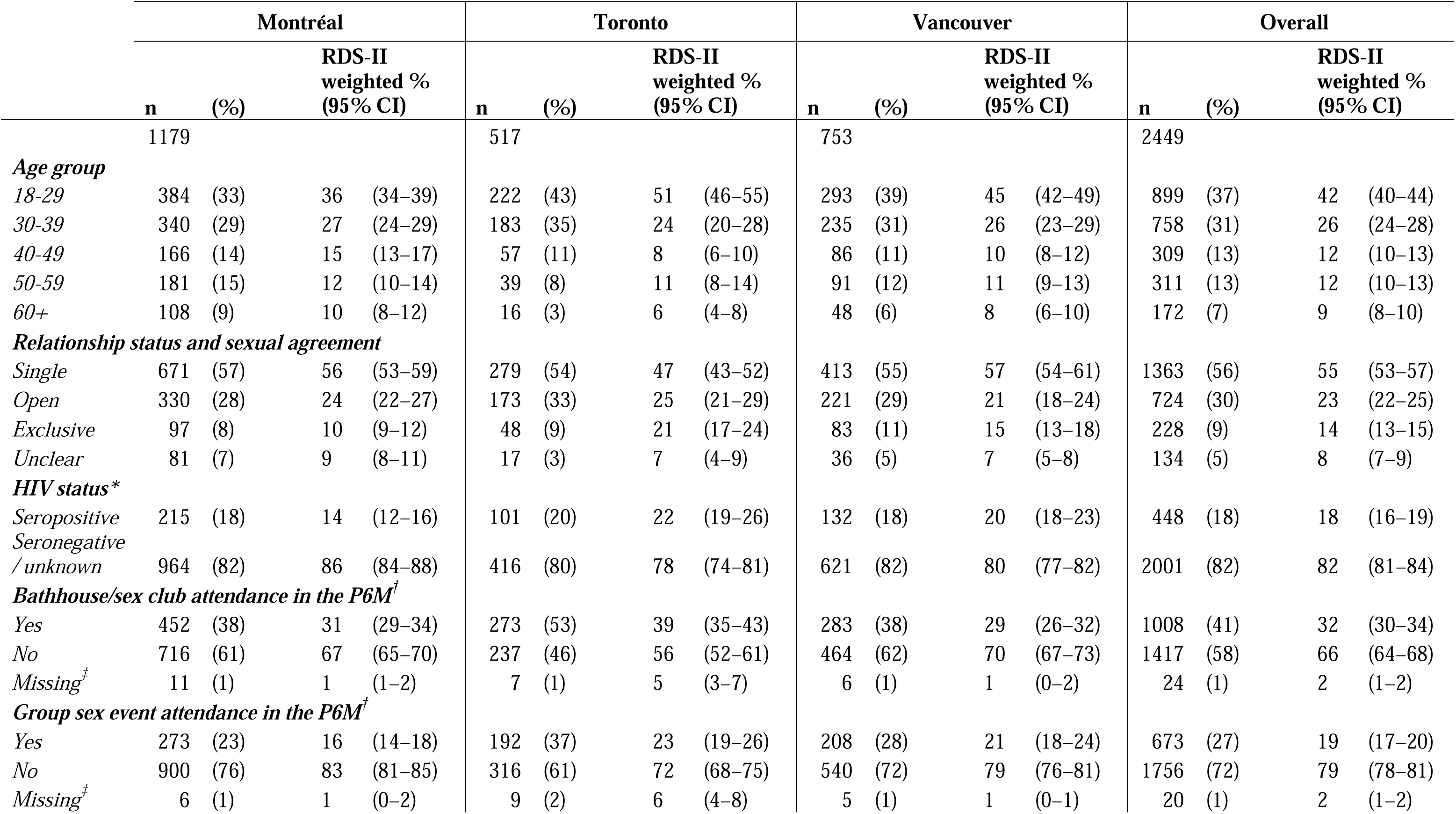
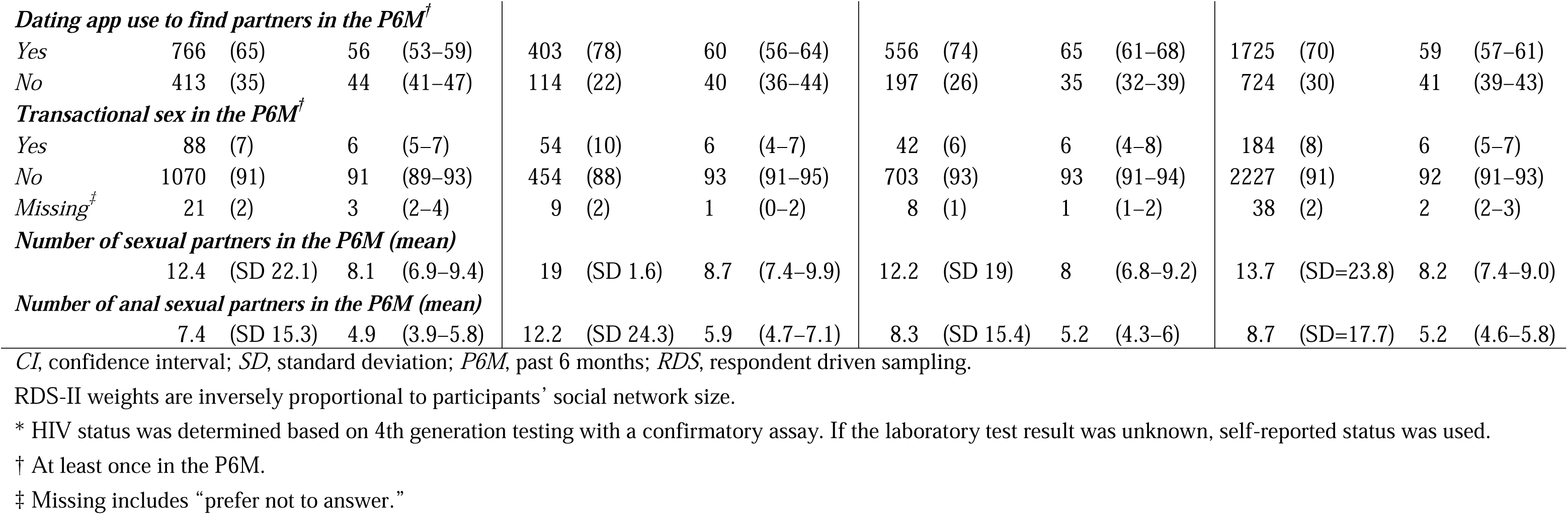
Unadjusted and RDS-II adjusted baseline estimates of the number of sexual partners in the past six months and its correlates among the *Engage Cohort Study* participants in Montréal, Toronto, and Vancouver, 2017–2019.

### Correlates of the networks’ number of sexual partners

In all three cities, a higher number of sexual partners in the post-restrictions period was strongly associated with attendance of group sex events, with a rate ratio (RR) of 3.65 (95% Credible Interval [CrI]: 2.79–4.74) in Montréal, 3.44 (95%CrI: 2.43–4.83) in Toronto, and 3.08 (95%CrI: 2.22–4.32) in Vancouver. Other strong correlates were participation in transactional sex, usage of dating apps, and visit to bathhouses and/or sex clubs (***Table S3***).

### Differences in the distribution of sexual partner numbers by city and time period

Overall, the fitted distribution of sexual partner numbers was similar across the three cities: the mean number of partners was 10.3 (95%CrI: 9.3-11.3) in Montréal, 12.8 (95%CrI: 11.1-14.7) in Toronto and 10.6 (95%CrI: 9.41-11.9) in Vancouver. However, pre-pandemic, sexual networks in Toronto had the heaviest-tailed distribution, with 1.2% (95%CrI: 0.8-1.6%) of GBM reporting ≥100 partners in the P6M, compared with 0.5% (95%CrI: 0.3-0.7%) in Montréal and 0.3% (95%CrI: 0.1-0.5%) in Vancouver. All posterior distribution samples showed that Toronto had a larger cumulative density of ≥100 numbers of sexual partners than Montréal and Vancouver. This result held during the post-restrictions period: 0.5% (95%CrI: 0.2-0.9%) of GBM in Toronto reported ≥100 partners, 0.3% (95%CrI: 0.2-0.5%) in Montréal, and 0.4% (95%CrI: 0.2-0.7%) in Vancouver. Post-restrictions, Toronto had a larger cumulative density of ≥100 numbers compared to Montréal and Vancouver (83% and 63% of the posterior distribution samples, respectively; ***Figure 1; Figure S1***).

**Figure 1.**
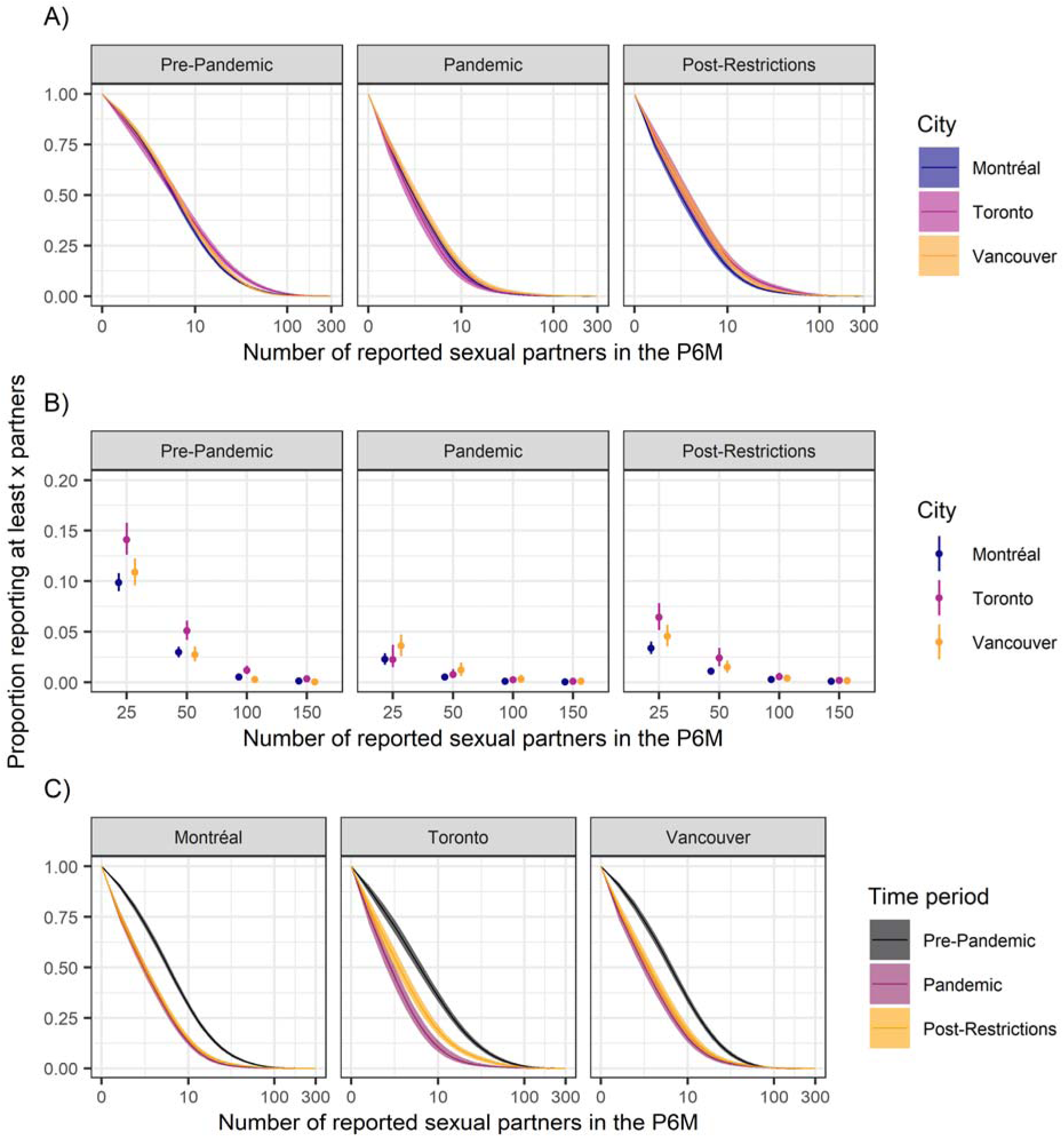
Cumulative distribution of sexual partner numbers in the past 6 months across Montréal, Toronto, and Vancouver at each time period, weighted by respondent driven sampling (RDS-II) and inverse probability of censoring weights, with 95% credible intervals. A) Full distribution (one time period per panel), B) Selected values (one time period per panel), C) Full distribution (one city per panel). P6M: past 6 months.

Compared to the pre-pandemic period, all three cities witnessed a marked reduction in the distribution of sexual partner numbers during the COVID-19 pandemic. In all three cities across all 4,000 posterior distribution samples, the cumulative density of ≥25 sexual partners in the P6M were consistently larger for the pre-pandemic vs. pandemic period. Sexual activities appeared to have rebounded after lifting travel restrictions: in Montréal, Toronto, and Vancouver, 100%, 100%, and 92% of posterior distribution samples showed a larger proportion of participants reporting ≥25 sexual partners in the P6M as compared to the pandemic period. However, sexual activities have not fully recovered to pre-pandemic levels: in all three cities, 100% of the posterior distribution samples had a greater proportion of GBM with ≥25 sexual partners in the pre-pandemic than the post-restrictions period (***Figure 1***).

### Basic reproduction numbers from reported case counts and the next-generation matrix

Based on the reported cases counts, we estimated an *R_0_*of 2.55, 2.53 and 2.44 in Montréal, Toronto and Vancouver, respectively (***Figure 2A***). We also estimated a cumulative incidence proportion of mpox-diagnosed GBM ranging from 0.6–0.9% in all cities, assuming that GBM comprise 2.9% of all men ≥15 years old in each city (***Table S5***, ***Figure S2***).

**Figure 2.**
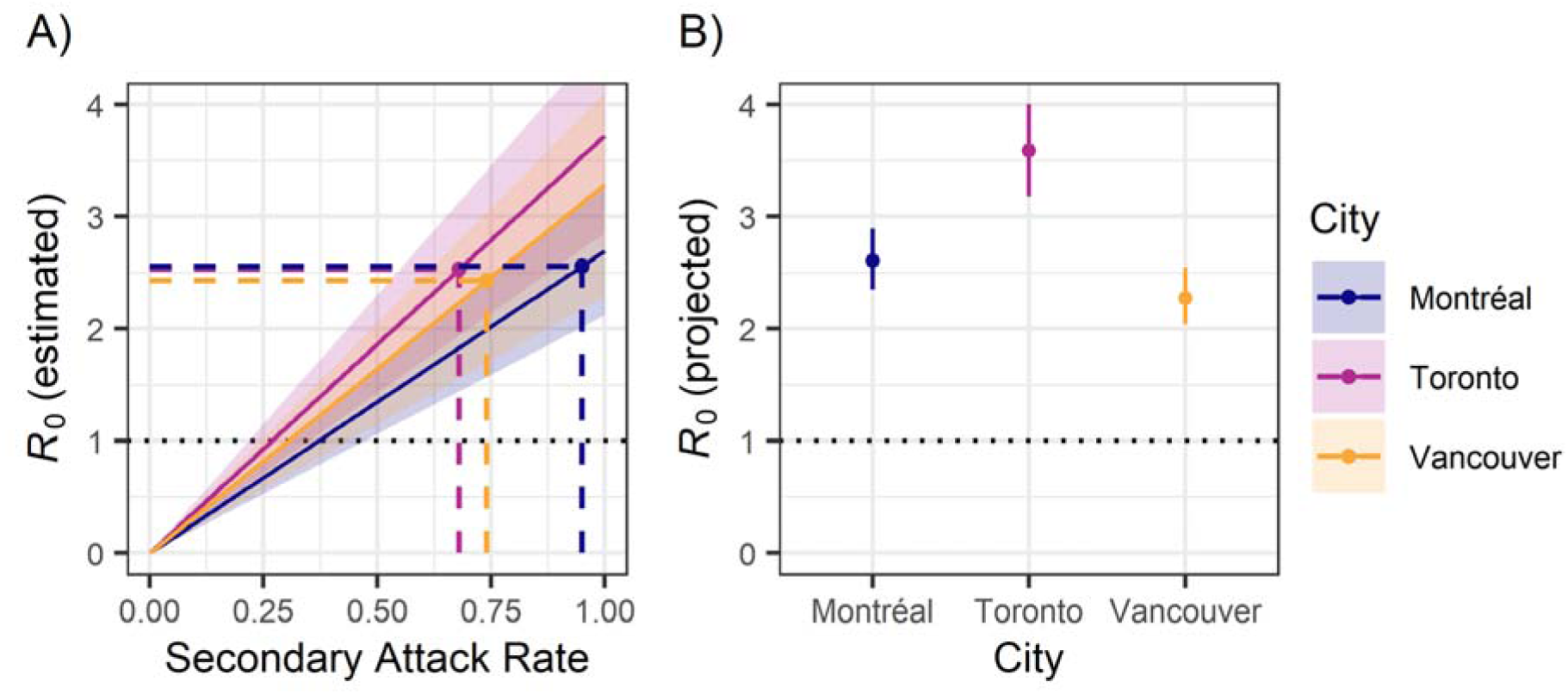
Basic reproduction number (*R_0_*) of mpox. A) City-specific *R_0_* estimates based on the initial growth rate of reported mpox cases (dashed lines) and on the fitted distribution of sexual partner numbers using the next-generation matrix method in the post-restrictions period (solid lines, shaded area: 95% credible intervals). B) Projected *R_0_*to the pre-pandemic period, assuming pre-pandemic sexual activity and using the mean of the three cities’ secondary attack rate (79%). Next-generation matrix assumes proportionate mixing of sexual activity groups is assumed. P6M: past 6 months.

By comparing the *R_0_* based on mpox cases to the NGM *R_0_*estimated from the partnership distributions, we estimated that the average per-partnership SAR across cities was 79%. Based on this SAR, had the distribution of sexual partner numbers been at pre-pandemic levels, the mpox *R_0_* using the NGM would have been 2.61 (Montréal), 3.59 (Toronto), and 2.27 (Vancouver; ***Figure 2B***).

### Sensitivity analyses

When restricting the sample to only participants with follow-up at each time period, the distributions of sexual partner numbers were broadly similar for all cities and time periods (***Figure S3***). Similarly, standardizing the distributions to the covariate distribution in Montréal did not substantially change the results for Vancouver. However, in Toronto, the tail of the distribution of sexual partner numbers was slightly lighter in Toronto after standardization, especially for the pre-pandemic and post-restrictions time periods (***Figure S4***). When using anal sexual partner as outcome, the comparisons across cities and time periods did not qualitatively change, but the distributions had smaller means and lighter tails (***Figure S5***).

Overall, these sensitivity analyses suggest that the results are relatively robust to the weighting methods and to differences in covariate distributions across the cities. However, the expected *R_0_* and SAR would be lower based on anal sexual partners as compared to all sexual partners.

## Discussion

In a large RDS cohort of GBM in Canada’s three largest cities, there was a marked decrease in the distribution of sexual partner numbers during the COVID-19 pandemic compared to the pre-pandemic period (2017–2019). Despite a small increase after travel restrictions were lifted (late 2021–early 2023), GBM sexual activity was well below pre-pandemic levels at the time of the 2022–2023 mpox outbreak. Despite the reductions in sexual partnerships, the *R_0_* of mpox was approximately 2.5 in all three cities during the 2022–2023 mpox outbreak. As the population’s social and sexual behaviors are expected to recover towards pre-pandemic levels, the mpox *R_0_* may increase; it is therefore essential to continue public health surveillance and preventative activities —community outreach, vaccination— to mitigate the local impacts of mpox re-introductions into Canada.

We found that GBM had substantially fewer sexual partners in all three cities during the COVID-19 pandemic, and sexual activity remained lower than pre-pandemic levels even after restrictions were lifted. These findings are in line with previous research from Canada^45,46^ and Europe^19,47,48^ which suggest that GBM sexual behaviours were influenced by public health measures and messaging related to the COVID-19 pandemic. The implications of our findings are that as sexual behaviours are expected to return towards pre-pandemic levels, future mpox outbreaks remain possible. Outbreak risks are also further amplified with case importation risks in an interconnected world and limited availability of mpox vaccines and therapeutics in countries in Africa where mpox has been endemic for decades.^49^

We found that attendance of group sex events, participation in transactional sex, usage of dating apps, and visits to bathhouse and/or sex clubs were associated with higher numbers of sexual partners in urban Canadian GBM. Notably, group sex events, use of dating apps, and visits to bathhouse and/or sex clubs were also associated with earlier mpox cases during the 2022-2023 outbreak.^2,12^ In the context of ongoing low coverage of 2^nd^ dose mpox vaccination across the three cities, and across other cities in Canada, these venues therefore provide a potentially interesting focus for prioritized and tailored vaccine strategies to increase coverage. For example, pop-up vaccine clinics could be set at bathhouses and sex clubs in partnership with community organizations.^50–52^ Additionally, focused communication campaigns to promote mpox vaccination could be rolled out on GBM-dating apps.^53,54^

Across Montréal, Toronto, and Vancouver, we estimated an *R_0_*of about 2.5 based on the incidence data. This is comparable to the 2.4 observed in Italy and in a pooled analysis of data from European countries.^55,56^ However, there is high variability in *R_0_*estimates from the pooled European analysis, with estimates ranging from 1.6 (Belgium) to 3.0 (Italy).^56^ Despite this high *R_0_*, we estimated a cumulative incidence proportion of only 0.6-0.9% of GBM by October 2022. Similarly, an analysis by Murayama et al.^15^ found that epidemic growth reached its peak at cumulative incidence proportions of 0.15–0.47% in various North American and European countries. Such estimates are substantially lower than the theoretical “herd immunity threshold” of 60% (assuming homogeneous contact rates) implied by the current *R_0_* estimates, although *R_0_* < 1 may be more readily achieved if those at higher risk are disproportionately protected by vaccination and/or prior infection.^57^

The results should be interpreted considering three main limitations. First, our post-restrictions period overlaps with the time when spread of the SARS-CoV-2 Omicron variant took place. Thus, the estimated distribution of sexual partner numbers may have been affected by measures introduced in response to the Omicron SARS-CoV-2 wave. However, these restrictions were relatively short-lived, and our definition enabled consistent and comparable time period definitions across cities.^58^ Second, although we used IPCW to address attrition bias, this bias may not have been fully adjusted if the loss to follow-up model was misspecified (i.e., not all variables associated with attrition were included). Lastly, our quantification of mpox transmission potential assumed proportionate (to degree) mixing among sexual activity groups for computational simplicity. If mixing was “like-with-like” (assortative) by sexual activity, the modeled *R_0_*given SAR would increase, and thus the inferred SAR would decrease.

Our approach to estimating the distribution of sexual partner numbers has several strengths. First, we implemented both RDS-II weights and IPCW to obtain estimates representative of sexually active GBM in the three largest Canadian cities. Furthermore, inter-city comparisons enabled us to account for potential differences in GBM communities in each city and explore their relative impact on the transmission dynamics of mpox. Finally, *Engage*’s longitudinal data collection allowed us to quantify behavioural changes among GBM from pre-COVID-19 pandemic up to February 2023.

## Conclusion

In Montréal, Toronto, and Vancouver, GBM had fewer sexual partners during the COVID-19 pandemic. Even after travel restrictions were lifted in late 2021, sexual activities among urban Canadian GBM had not fully recovered to pre-pandemic levels. The overall distribution of sexual partner numbers was similar across cities, potentially explaining the similar observed cumulative fraction of mpox cases among GBM in the three cities. With sexual activity expected to return to pre-pandemic levels, public health authorities should consider the potential risk of mpox resurgence. Improving first- and second-dose vaccination coverage among individuals at risk with high numbers of sexual partners should be prioritized.

## Data Availability

All data produced in the present study are available upon reasonable request to the *Engage Cohort Study*.

## Acknowledgments

The authors would like to thank the *Engage* study participants, office staff, and community engagement committee members, as well as our community partner agency, *RÉZO*. Engage/Momentum II is funded by the Canadian Institutes for Health Research (CIHR, #TE2-138299), the CIHR Canadian HIV/AIDS Trials Network (#CTN300), the Canadian Foundation for AIDS Research (CANFAR, #Engage), the Ontario HIV Treatment Network (OHTN, #1051), the Public Health Agency of Canada (Ref: 4500370314), Toronto Metropolitan University, Canadian Blood Services (#MSM2017LP-OD), and the Ministère de la Santé et des Services sociaux (MSSS) du Québec.

## Declarations

### Author contribution

JLFA, FX, and MM-G contributed to the conception and design. JC, DG, TAH, MD, and SM were involved in the design, data collection, and data management of the *Engage Cohort Study*. Analyses were performed by JLFA and FX, with support from MM-G. JK, LW, OG, HS, MI, and SM provided input on preliminary methods discussions. The manuscript was drafted by FX and JLFA. All authors contributed to the interpretation of results and reviewed the manuscript for important intellectual content. Overall supervision for this project was provided by MM-G. All authors approved the final manuscript.

### Software and source code

Analysis code is available on a *GitHub* repository (https://github.com/pop-health-mod/mpox-engage-sex-networks).

