## Supplementary Materials for "Characteristics of the sexual networks of gay, bisexual, and other men who have sex with men in Montréal, Toronto, and Vancouver: implications for the transmission and control of mpox in Canada"

### Table of contents

Supplementary Methods

Supplementary Tables S1 to S5

Supplementary Figures S1 to S6

References

**Supplementary Methods**

**Table S1. Definitions of analysis variables.**

| Notation | Definition | Question from the Engage Cohort Study |
|---|---|---|
| **y** | Number of all-type sexual partners in the past 6 month (P6M)<br>Number of anal sexual partners in the P6M (for ***Sensitivity Analysis***) | 5.5 During the PAST 6 MONTHS, with how many guys have you had any kind of sex (anal, oral, mutual masturbation, rimming, frontal/vaginal, etc.)?<br>_____ guys<br>5.12 During the past 6 months, how many guys have you had anal sex with (as top or bottom)? _____ guys |
| **n** | Number of Participants | See ***Table S2*** for the sample size at each city-timepoint |
| **network size** | Participant network size (for RDS-II estimation) | How many men who have sex with men aged 16 years or older, including trans men, do you know who live or work in the [Metro Vancouver/Greater Toronto/Metro Montreal depending on site] area (whether they identify as gay or otherwise)? This includes gay/bi guys you see or speak to regularly; e.g., close friends, boyfriends, spouses, regular sex partners, roommates, relatives, people you regularly hang out with, etc. |
| ***Age*** | Age at time of the visit | 1.3 What is your age (i.e., how old are you)? |
| ***RelationshipStatus*** | Relationship status at time of the visit | 2.18 Do you currently have a relationship with a main partner? No\|Yes<br>2.23 What discussions have you and your main partner had with each other in terms of only having sex with each other?<br>We haven't explicitly discussed only having sex with each other or not\|We have discussed only having sex with each other, but have not agreed to anything\|We have discussed only having sex with each other and agreed to only have sex with each other\|We agreed to have other sex partners, but only ones we share (we only play together)\|We agreed to have other sex partners, some of whom we share and others whom we see separately (we play together and separately)\|We agreed to have other sex partners whom we only see separately (we only play separately)\|We agreed to another arrangement. Please describe: __\|No main relationship partner |
| ***HIVStatus*** | HIV serostatus | Derived by *Engage Cohort Study* from participant laboratory-tested and |

| | | |
|---|---|---|
| | | self-report HIV serostatus |
| ***Bathhouse*** | Visit to bathhouses and/or sex clubs during the P6M | 5.46 During the past 6 months did you go to a bathhouse or sex club?<br>No\|Yes\|Don’t know / don’t remember\|Prefer not to answer |
| ***Groupsex*** | Attendance in group sex events during the P6M | 5.47 During the past 6 months did you attend any group sex events?<br>By group sex we mean sex where 4 or more people get together and have some kind of sex with some or all of the other people there. This could include at a private organized sex party, at a bathhouse, in darkrooms, or other venues.<br>No\|Yes\|Don’t know / don’t remember |
| ***DatingApp*** | Dating app usage during the P6M | 4.3 In the PAST 6 MONTHS have you used a smartphone app or internet website to connect with other guys?<br>Never\|Less than once per month\|About once per month\|More than once per month\|Prefer not to answer<br><br>Note: this variable was only reported at the baseline (Pre-Pandemic) |
| ***TransactionalSex*** | Participation in transactional sex (received money or goods in exchange for sex) in the P6M | 5.45 In the past 6 months, have you...<br>(Remember that for the following question, by “sex” we mean oral sex, anal sex, frontal/vaginal sex, masturbation, rimming, fisting, sex toys, or watersports.)<br>i. RECEIVED money in exchange for sex? h. RECEIVED drugs in exchange for sex? m. RECEIVED other goods or services (e.g.,room, meal, gifts) in exchange for sex?<br>Yes\|No\|Don’t know / Don’t remember\|Prefer not to answer |

***Distribution of sexual partner numbers (Bayesian regression and post-stratification)***

Our main approach to estimate the distribution of sexual partner numbers consisted of three steps. First, we fit a negative binomial regression model to the reported number of sexual partners in the past 6 months (P6M). The model covariates were age group, relationship status and sexual arrangement, HIV serostatus, visit to bathhouses and/or sex clubs, attendance to group sex events, use of dating apps, and participation in transactional sex (***Table S1***). For each city-time period $j$, we fit this regression model, and then obtained the fitted posterior distribution of the mean number of sexual partners for each participant $i$, i.e., $\mathbb{E}[\hat{y}_{i,j}|X_{i,j}]^{(m)}$.

The regression model can be written as

$$\log \mathbb{E}[y] = \alpha_j + \beta_j X_j$$

where

$i :=$ index for participants, $i \in \{1,2,\ldots,n_j\}$;

$j :=$ city and time period, $j \in$ {*Montréal-Pre-Pandemic, Toronto-Pre-Pandemic, Vancouver-Pre-Pandemic, Montréal-Pandemic, Toronto-Pandemic, Vancouver-Pandemic, Montréal-Post-Restrictions, Toronto-Post-Restrictions, Vancouver-Post-Restrictions*};

$X_j :=$ an $i \times j$-dimension matrix containing the values of predictors for all participants for city and time period $j$, i.e., *Age, RelationshipStatus, HIVStatus, Bathhouse, Groupsex, DatingApp, TransactionalSex*;

$y :=$ number of all-type sexual partners in the P6M;

$m :=$ index of samples from the posterior, $m \in \{1,2,\ldots,4000\}$;

$\alpha_j :=$ model intercept for city-time period $j$; and

$\beta_j :=$ vector of regression coefficients for city-time period $j$, for set of covariates $X_j$.

We used non-informative priors for the negative binomial regressions. We used a $Normal(0, 10)$ distribution for the regression intercept and all predictor coefficients. For the over-dispersion parameter $\phi$, we used a $halfCauchy(0, 5)$ distribution.

Second, in each city-time period $j$, for each participant $i$, we used the participant's posterior predictive mean $\mathbb{E}[\hat{y}_{i,j}|X_{i,j}]$ and the overdispersion parameter $\phi$ to compute the probability of observing $k$ partners for that participant, where $k \in \{1,2,\ldots,300\}$ (the largest number of reported partners in the P6M at baseline was 300). We performed this procedure separately for all $m$ samples.

Third, we performed post-stratification to incorporate respondent-driven sampling (RDS)-II weights and inverse probability of censoring weights (IPCWs), to adjust for the RDS sampling design and loss to follow-up, respectively. These weights were used to estimate a more representative distribution of sexual partner numbers in the P6M, the RDS-IPC-weighted distribution (computation of RDS-II weights, IPCWs, and the RDS-IPC weights is explained in the next section). The distribution of sexual partner numbers can be thought of as the proportion of men who report $k$ partners in the P6M, i.e., $P(y_j = k)$ for $k \in \{1,2, \dots ,300\}$. For each city-time period, the RDS-IPC adjusted distribution of partner numbers can thus be estimated using the equation

$$P(y_j = k)^{(m)} = \sum_{i=1}^{n_j} \frac{P\left(y_j = k | \mathbb{E}[\hat{y}_{i,j} | X_{i,j}]^{(m)}\right) w_{i,j}}{\sum_{i=1}^{n_j} w_{i,j}}$$

where individual probabilities $P(y_{j,i} = k)$ are computed from each participant's posterior predictive mean from the second step, and $w_{j,i}$ is the RDS-IPC weight for participant $i$ at city and time period $j$. We used the mean of the posterior distribution as the point estimate and computed 95% credible intervals (CrI) from the 2.5th and 97.5th percentiles.

We verified that the data satisfies the assumptions for fitting a negative binomial regression. Briefly, we verified that the independence, linearity, and overdispersion assumption of the negative binomial regression model were satisfied. Further, the percentage of the population reporting 0 partners was around 13% in each city during the pandemic and post-restrictions timepoints, and therefore we chose not to use a zero-inflated model.

***RDS-II weights and inverse probability of censoring weights***

We computed RDS-II weights for each city separately, and the inverse probability of censoring weights (IPCW) for the two follow-up time periods (separately for each city). The RDS-II weights were computed using the RDS-II estimator and the self-reported network size, capped at 150 (a correction was applied if a participant reported knowing less gay men than they had recruited). For a participant $i$, the RDS-II weight was

$$\widetilde{w}_{RDS}^{l,i} = \frac{\sum_{i=1}^{n_l} network\ size_i}{n_l} \frac{1}{network\ size_i}$$

and the normalized RDS-II weight was

$$w_{RDS}^{l,i} = \widetilde{w}_{RDS}^{l,i} \frac{n_l}{\sum_{i=1}^{n_l} \widetilde{w}_{RDS}^{l,i}}$$

where $l \in \{$ *Montréal, Toronto, Vancouver* $\}$.

For IPCW, we computed the propensity score for being loss to follow-up (LTFU),

$P(LTFU = 1)$. For the pandemic and post-restrictions periods, a participant was considered LTFU if they did not have a visit during the defined time period. We identified potential predictors of LTFU by computing RDS-weighted standardized mean differences (SMD) to assess the imbalance in the predictors (measured at pre-pandemic) between LTFU and retained participants. All identified LTFU predictors (i.e., with imbalance as measured by SMD) were used in the propensity score model in matrix $Z$ (variable definitions in ***Table S1***):

$$logit\left(P(LTFU = 1)_j\right) = \alpha'_j + \beta'_j Z_j$$

where

$Z :=$ an $i \times j$-dimension matrix containing the values of predictors for all participants for city and time period $j$, i.e., *Age, RelationshipStatus, HIVStatus, Bathhouse, Groupsex, DatingApp, TransactionalSex*;
$\alpha'_j :=$ model intercept for city-time period $j$; and
$\beta'_j :=$ vector of regression coefficients for city-time period $j$, for set of covariates $X_j$.

The propensity score $ps$ for participant $i$ being lost to follow at time period $j$ is therefore $ps = P\left(LTFU = 1|Z_{i,j}\right)$. The derived stabilized RDS-IPC weight for participant $i$ is

$$w_{i,j} = \begin{cases} w_{RDS}^{l,i} & \text{for the pre} - \text{pandemic time period,} \\ w_{RDS}^{l,i}\left(\dfrac{1}{1-ps}\right)\left(1 - \dfrac{w_{RDS}^{l,i} I(LTFU_{i,j} = 1)}{n_j}\right) & \text{if } LTFU_{i,j} = 0, \text{and} \\ w_{RDS}^{l,i}\left(\dfrac{1}{ps}\right)\left(\dfrac{w_{RDS}^{l,i} I(LTFU_{i,j} = 1)}{n_j}\right) & \text{otherwise.} \end{cases}$$

Finally, we ensured that the RDS-IPC weights sum to the RDS-adjusted number of participants, i.e., $\sum_{i=1}^{n_j} w_{i,j} = \sum_{i=1}^{n_j} w_{RDS}^{l,i}$ and that the SMD after the adjustment is small.

***Reproduction number from the distribution of sexual partner numbers using the next-generation matrix***

We constructed the transmission ($T$), transition ($\Sigma$) and auxiliary ($E$) matrices following Diekmann and colleagues,[3] under a simplified Susceptible-Exposed-Infectious-Removed model, assuming no interventions or behavioural changes during the epidemic. Model parameters are presented in ***Table S4*** and the model equations are:

$$\begin{cases} \frac{\partial S_i}{\partial t} = -\lambda_i S_i \\ \frac{\partial E_i}{\partial t} = \lambda_i S_i - v E_i \\ \frac{\partial I_i}{\partial t} = v E_i - \gamma I_i \\ \frac{\partial R_i}{\partial t} = \gamma I_i \end{cases}$$

where

$i :=$ sexual activity group for $i \in \{1,2,\ldots,300\}$;

$\beta :=$ secondary attack rate (SAR);

$\lambda_i :=$ force of infection for the $i$-th sexual activity group, $\lambda = c_i \beta \frac{\sum_{j=1}^{100} c_j I_j}{\sum_{j=1}^{100} c_j N_j}$;

$c_i :=$ average degree of sexual partners in the P6M in the $i$-th sexual activity group;

$v :=$ rate of transition from exposed to infectious (incubation period)$^{-1}$; and

$\gamma :=$ rate of recovery, from infectious to recovered/removed (infectious duration)$^{-1}$.

At $t = 0$, $E_i = 0, S_i = N_i \forall i$, we can therefore simplify the force of infection as

$$\frac{\partial E_i}{\partial t} = \lambda_i S_i = c_i \beta \frac{\sum_{j=1}^{100} c_j I_j}{\sum_{j=1}^{100} c_j N_j} N_i = c_i \beta \frac{d_i}{\sum_{j=1}^{100} c_j d_j} \sum_{j=1}^{100} c_j I_j$$

where

$N_i :=$ size of the $i$-th sexual activity group; and

$d_i :=$ density (proportion) of the $i$-th sexual activity group in the population.

Therefore, the matrices $T$, $\Sigma$ and $E$ can be expressed

$$\boldsymbol{T_{200*200}} = \begin{cases} \frac{c_{\frac{i+1}{2}} c_{\frac{j}{2}} d_{\frac{i+1}{2}} \beta}{\sum_{l=1}^{100} c_l d_l} & \text{for } i = 1, 3, \ldots, 199 \wedge j = i + 1 \\ 0 & \text{otherwise} \end{cases}$$

$$\boldsymbol{\Sigma}_{\mathbf{200*200}} = \begin{cases} -v & \text{for } i = 1,3,...,199 \wedge i = j \\ -\gamma & \text{for } i = 2,4,...,200 \wedge i = j \\ v & \text{for } i = 2,4,...,200 \wedge j = i-1 \\ 0 & \text{otherwise} \end{cases}$$

$$\boldsymbol{E}_{\mathbf{200*100}} = \begin{cases} 1 & \text{for } i = 1,3,...,199 \wedge j = \frac{i+1}{2} \\ 0 & \text{otherwise} \end{cases}$$

where $i$ and $j$ are the row and column indices, respectively.

These three matrices allowed for the construction of a next-generation matrix, and $R_0$ was obtained from the largest non-zero eigenvalue of the matrix. We repeated the procedure for each city at the post-restriction time period. Given the uncertainty in the SAR $\beta$ for mpox, we computed $R_0$ for a range of values for $\beta$ between 0 and 1. We then estimated a plausible SAR by combining these estimates with the $R_0$ estimated from growth rate in cases (explained below), and used this SAR value to project the $R_0$ based on the pre-pandemic distribution of sexual partner numbers.

***Reproduction number from reported case numbers and endpoint of exponential growth period***

We also estimated the $R_0$ from the cumulative incidence of confirmed mpox cases in the three provinces where Montréal, Toronto, and Vancouver are located (respectively, Québec, Ontario, and British Columbia), using the formula $R_0 = 1 + D\Lambda$ where $D$ is the duration of infectiousness (17.3 days) and $\Lambda$ is the epidemic growth rate.

To estimate $\Lambda$, we used the period in the outbreak during which mpox cases were growing exponentially. We ascertained the end of the exponential growth period by visual inspection of the curve of cumulative cases and by determining the initial period during which the effective reproductive number ($R_t$) was relatively stable. We estimated the $R_t$ from the reported cases using the *EpiEstim* package,[9] using a 14-day smoothing window and focusing on the first 200 days of the outbreak in each city, when the majority of mpox cases were diagnosed (Québec: April 28th, 2022– November 14th, 2022; Ontario: May 13th, 2022–November 29th, 2022; British Columbia: May 25th, 2022–December 11th, 2022). Based on the curve of log-cumulative cases and the $R_t$, we estimated $\Lambda$ using the 50 days after the first mpox case was reported in a province (***Figure S2, S6***). Other natural history parameters were derived from literature and are summarized in ***Table S4***.

### Supplementary Tables

**Table S2. Retention of *Engage Cohort Study* participants at each time period.**

| | n (%) | | |
|---|---|---|---|
| | **Montréal** | **Toronto** | **Vancouver** |
| **Pre-Pandemic** | 1,179 (100%) | 517 (100%) | 753 (100%) |
| **Pandemic** | 831 (70%) | 302 (58%) | 449 (60%) |
| **Post-Restrictions** | 786 (67%) | 288 (56%) | 393 (52%) |

**Table S3. Association between number of sexual partners in the past 6 months and covariates among *Engage Cohort Study* participants in Montréal, Toronto, and Vancouver during the post-restrictions time period (December 2021–February 2023).**

| | Montréal | | Toronto | | Vancouver | |
|---|---|---|---|---|---|---|
| | **RR (95% CrI)** | **SE** | **RR (95% CrI)** | **SE** | **RR (95% CrI)** | **SE** |
| ***exp(intercept)*** | 4.08 (3.21, 5.22) | 0.0029 | 4.32 (3.08, 6.09) | 0.0042 | 4.38 (3.17, 6.19) | 0.0042 |
| ***Age group*** | | | | | | |
| ***18-29*** | REF | — | REF | — | REF | — |
| ***30-39*** | 1.17 (0.9, 1.52) | 0.0030 | 0.88 (0.61, 1.26) | 0.0043 | 1.26 (0.88, 1.76) | 0.0042 |
| ***40-49*** | 0.83 (0.61, 1.13) | 0.0033 | 0.82 (0.53, 1.25) | 0.0049 | 1.05 (0.69, 1.57) | 0.0046 |
| ***50-59*** | 0.70 (0.51, 0.98) | 0.0035 | 0.75 (0.41, 1.4) | 0.006 | 1.04 (0.68, 1.6) | 0.0048 |
| ***≥60*** | 0.72 (0.53, 0.98) | 0.0032 | 0.40 (0.23, 0.72) | 0.0057 | 1.05 (0.66, 1.65) | 0.0051 |
| ***Relationship status and sexual agreement*** | | | | | | |
| ***Single*** | REF | — | REF | — | REF | |
| ***Open*** | 1.11 (0.91, 1.36) | 0.0014 | 1.11 (0.82, 1.5) | 0.0025 | 0.94 (0.73, 1.21) | 0.0021 |
| ***Exclusive*** | 0.43 (0.32, 0.58) | 0.0022 | 0.47 (0.29, 0.78) | 0.0038 | 0.23 (0.15, 0.33) | 0.0033 |
| ***Unclear*** | 0.69 (0.49, 0.99) | 0.0024 | 0.43 (0.20, 0.97) | 0.006 | 0.88 (0.48, 1.67) | 0.0047 |
| ***HIV seropositive**** | 0.90 (0.71, 1.16) | 0.0019 | 1.21 (0.88, 1.65) | 0.0024 | 0.83 (0.6, 1.17) | 0.0031 |
| ***Bathhouse/sex club attendance in the P6M†*** | 1.73 (1.39, 2.16) | 0.0015 | 1.80 (1.32, 2.46) | 0.0028 | 1.80 (1.35, 2.43) | 0.0023 |
| ***Group sex event attendance in the P6M†*** | 3.64 (2.81, 4.72) | 0.0019 | 3.43 (2.43, 4.83) | 0.0032 | 3.06 (2.25, 4.16) | 0.0026 |
| ***Transactional sex in the P6M†*** | 3.63 (2.31, 5.98) | 0.0035 | 2.94 (1.79, 5.17) | 0.0041 | 3.69 (1.89, 8.20) | 0.0063 |
| ***1 / overdispersion parameter*** | 2.40 (2.18, 2.66) | 0.0007 | 3.08 (2.51, 3.88) | 0.0016 | 2.87 (2.42, 3.46) | 0.0015 |

Table presents the mean and 95% credible interval from 4,000 posterior samples from a Bayesian negative binomial regression model.

*CrI*, credible interval; *SE*, standard error; *RR*, rate ratio.

Dating app use was only evaluated at the baseline survey (pre-pandemic time period).

* HIV status was determined based on 4th generation testing with a confirmatory assay. If the laboratory test result was unknown, self-reported status was used.

† At least once in the P6M.

**Table S4. Natural history parameters for mpox.**

| Parameter (in days) | Symbol | Value* | (Range) | Sources |
|---|---|---|---|---|
| **Incubation period** | $v^{-1}$ | 7.9 | (7.5-9.0) | [10–12] |
| **Infectious duration** | $\gamma^{-1}$ or $D$ | 17.3 | (10-25) | [13–16] |
| **Mean of serial interval** | | 8.8 | (7.0-9.8) | [10,12,17] |
| **Standard deviation of serial interval** | | 8.7 | (4.2-10.9) | [10,12,17] |

* Values were estimated by averaging estimates reported by individual studies, weighted by study sample size.

**Table S5. Estimated cumulative incidence proportion of confirmed mpox cases among sexually active gay, bisexual, and other men who have sex with men during the 2022–2023 mpox outbreak.**

| | Montréal | Toronto | Vancouver |
|---|---|---|---|
| **Population size (all men ≥15 years old)*** | 1,735,065 | 2,529,370 | 1,102,200 |
| **Population size (sexually active GBM)*** | 50,317 | 73,352 | 31,964 |
| **Number of reported cases†** | 463 | 688 | 176 |
| **Cumulative incidence proportion** | 0.92% | 0.94% | 0.55% |

* Population size estimates were taken from the 2021 Canadian Population Census, for the corresponding census metropolitan area of each city. The population size of sexually active GBM was estimated as 2.9% of all men ≥15 years old.

† The number of confirmed mpox cases (as of October 7th, 2022) were reported from the *Public Health Agency of Canada*.

*GBM*, gay, bisexual, and other men who have sex with men.

**Supplementary Figures**

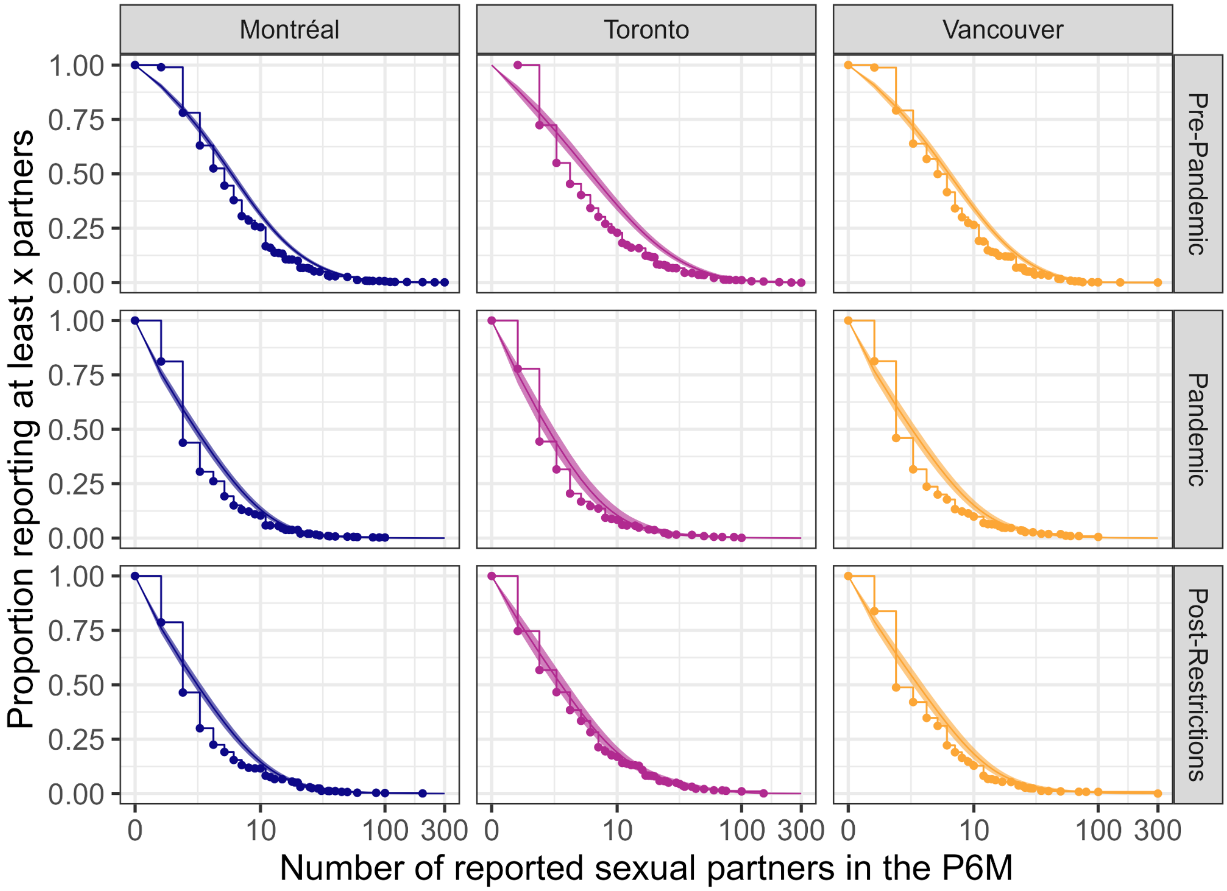


**Figure S1. Observed (RDS and inverse probability of censoring weighted) and fitted distributions of sexual partner numbers in the past 6 months for participants of the *Engage* cohort.** Lines with dots show the observed distributions, solid lines show the fitted distributions using negative binomial regression with post-stratification. Shaded area shows 95% credible intervals. RDS: respondent-driven sampling.

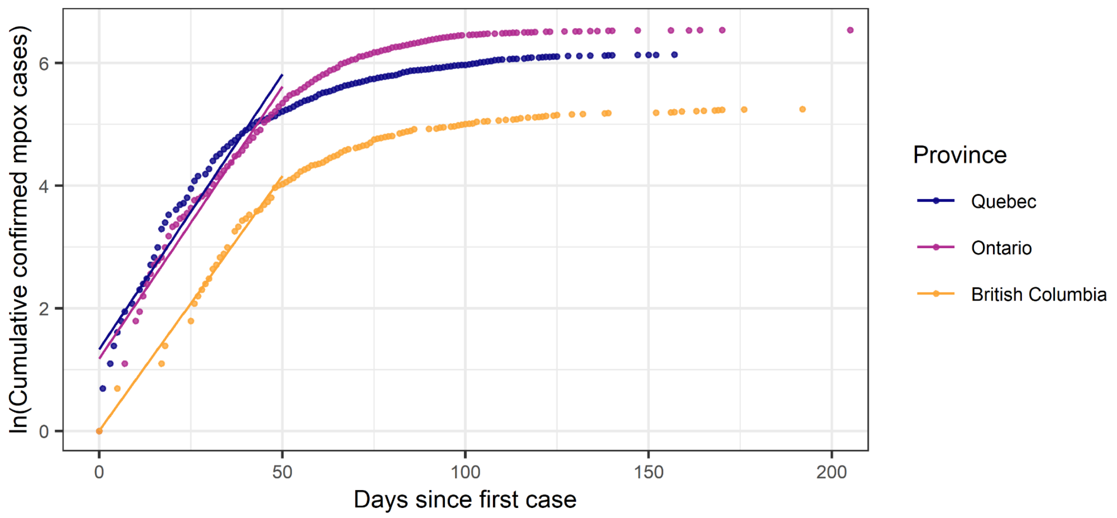


**Figure S2. Cumulative incidence of confirmed mpox cases in the provinces of Québec, Ontario, and British Columbia (natural log scale).** The growth rate used for estimating $R_0$ was computed as the slope of the log cumulative cases over time, using data from the first 50 days after the first mpox case was reported in each province (solid line shows the fitted regression).

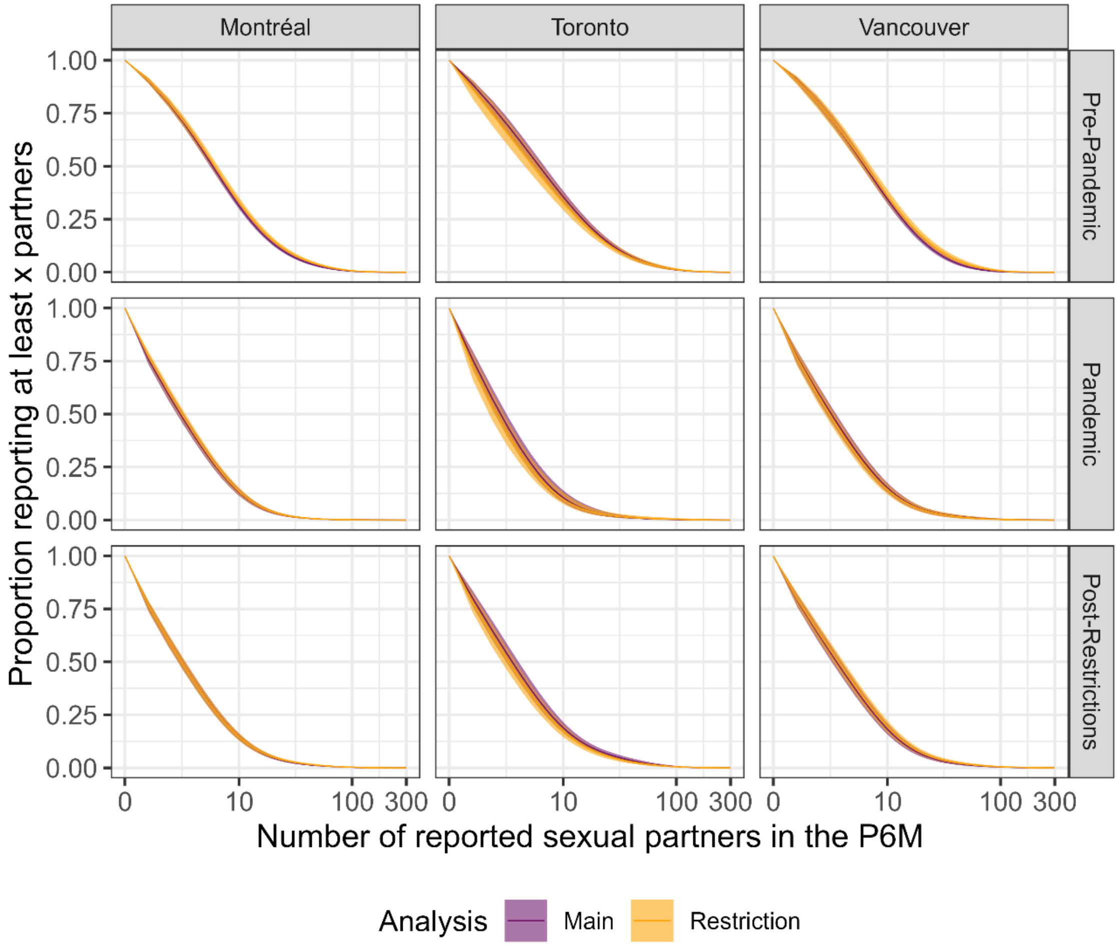


**Figure S3. Comparison of cumulative distribution of sexual partner numbers in the past 6 months between the main analysis (adjusted for RDS-IPC weights) and the restriction analysis (RDS-II weighted, using only participants with data for all time periods).** RDS: respondent-driven sampling; IPC: inverse probability of censoring.

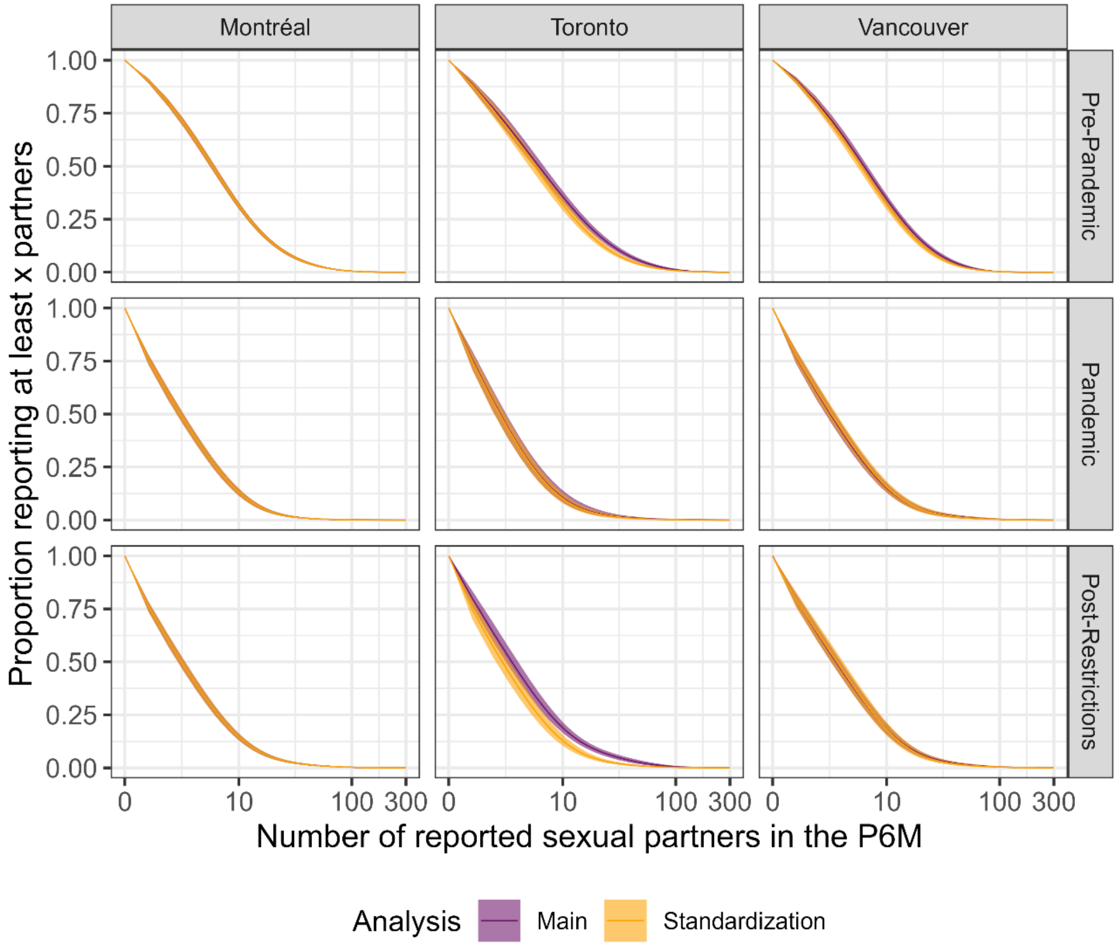


**Figure S4. Comparison of cumulative distribution of sexual partner numbers in the past 6 months between the main analysis (adjusted for RDS-IPC weights) and the standardization analysis (adjusted for RDS-IPC weights and standardized to the Montréal population).** RDS: respondent-driven sampling; IPC: inverse probability of censoring.

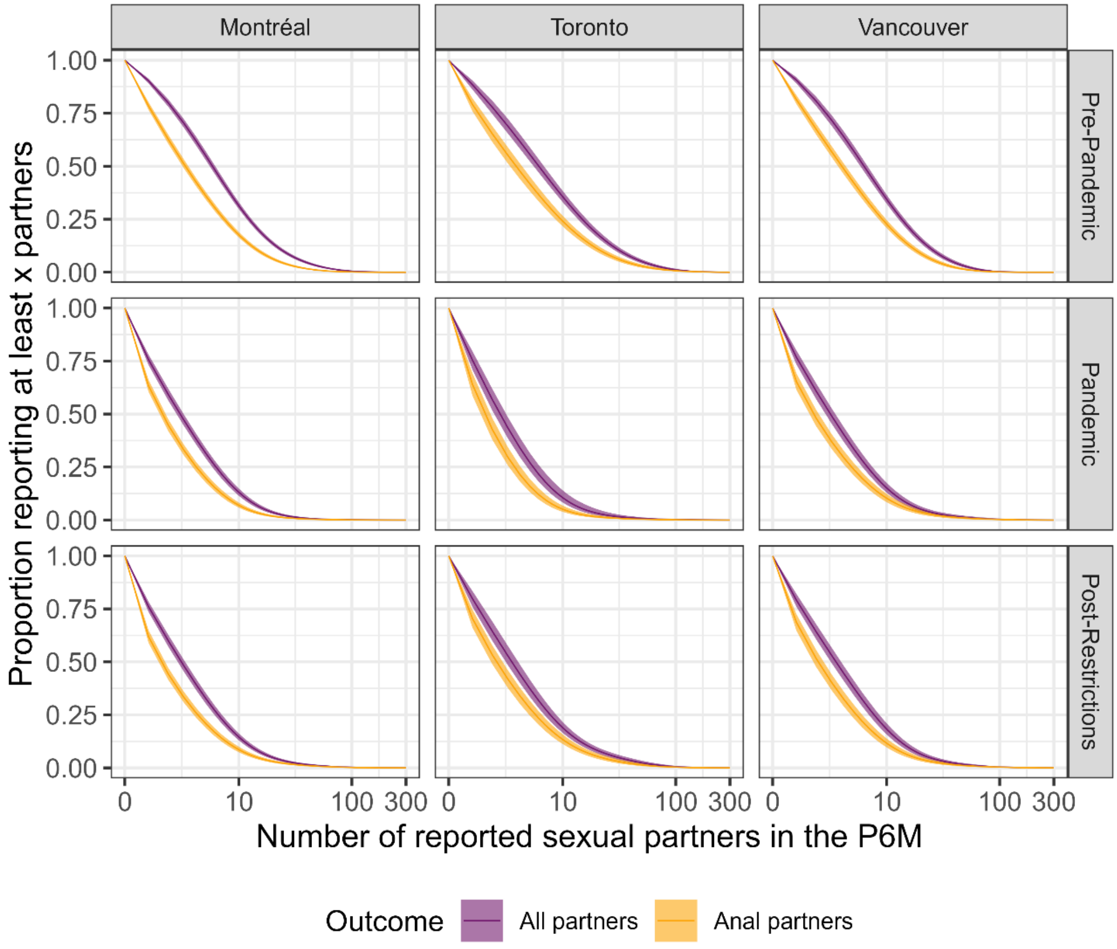


**Figure S5. Comparison of cumulative distribution of sexual partner numbers in the past 6 months between the main analysis (outcome: all sexual partners) and the sensitivity analysis using anal sexual partners as the outcome.** Both analyses were adjusted for RDS-IPC weights. RDS: respondent-driven sampling; IPC: inverse probability of censoring.

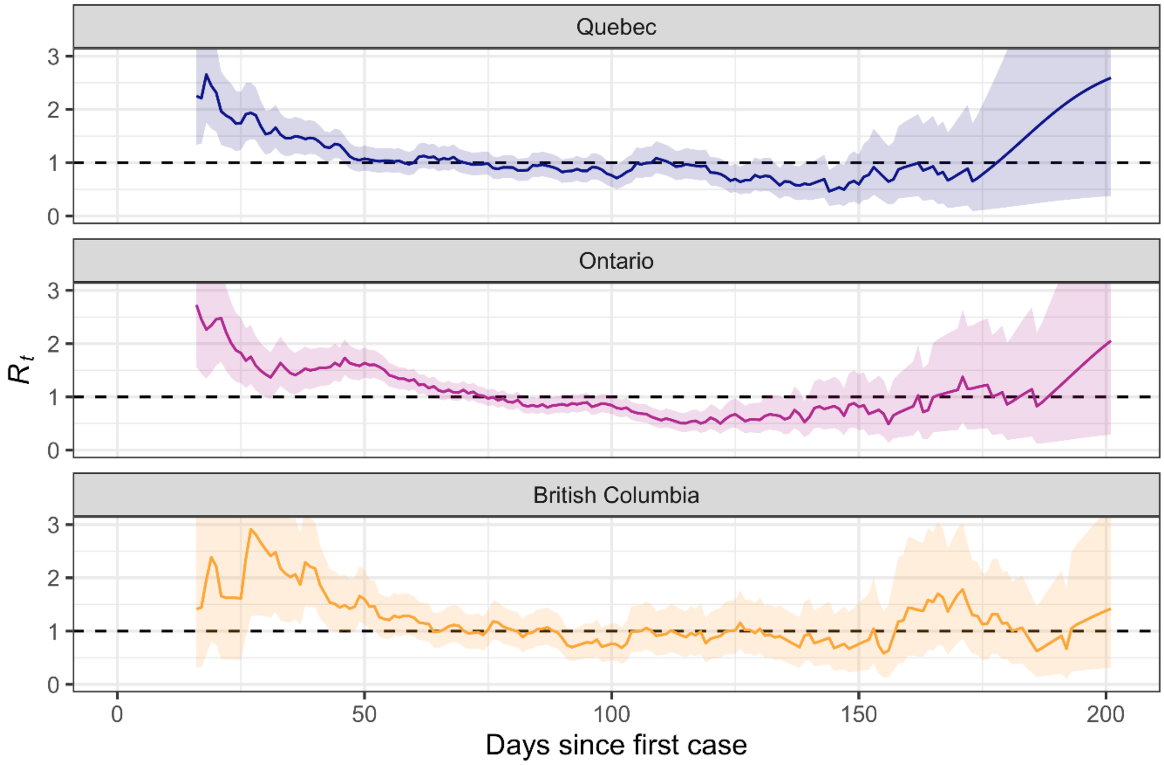


**Figure S6. Effective reproduction number ($R_t$) with 95% confidence interval in the province of Quebec, Ontario, and British Columbia.** $R_t$ was estimated based on confirmed mpox cases (data as of June 13th, 2023), using the first 200 days since the first case was reported in each province and a time window of 14 days**.**
